# Tryptophan metabolism, gut microbiota, and carotid artery plaque in women with and without HIV infection

**DOI:** 10.1101/2022.12.27.22283960

**Authors:** Kai Luo, Zheng Wang, Brandilyn A Peters, David B Hanna, Tao Wang, Christopher C Sollecito, Evan Grassi, Fanua Wiek, Lauren St Peter, Mykhaylo Usyk, Wendy S Post, Alan L Landay, Howard N Hodis, Kathleen M Weber, Audrey French, Elizabeth T Golub, Jason Lazar, Deborah Gustafson, Anjali Sharma, Kathryn Anastos, Clary B Clish, Rob Knight, Robert C Kaplan, Robert D Burk, Qibin Qi

## Abstract

**Background:** The perturbation of tryptophan (TRP)-kynurenine (KYN) metabolism has been linked with HIV infection and cardiovascular disease (CVD), but the interrelationship among TRP metabolites, gut microbiota, and atherosclerosis has not yet been fully understood in the context of HIV infection.

**Methods:** We included 361 women (241 HIV+, 120 HIV-) with carotid artery plaque assessments from the Women’s Interagency HIV Study, measured ten plasma TRP metabolites and profiled fecal gut microbiome. TRP metabolites related gut microbial features were selected through the Analysis of Compositions of Microbiomes with Bias Correction method. The associations of TRP metabolites and related microbial features with plaque were examined using multivariable logistic regression.

**Results:** While plasma kynurenic acid (KYNA) (odds ratio [OR]=1.93[1.12, 3.32] per one SD increase, *P*=0.02) and KYNA/TRP (OR=1.83[1.08, 3.09], *P*=0.02) were positively associated with plaque, indole-3-propionate (IPA) (OR=0.62 [0.40, 0.98], *P*=0.03) and IPA/KYNA (OR=0.51[0.33, 0.80], *P*<0.01) were inversely associated with plaque. Five gut bacterial genera and many affiliated species were positively associated with IPA (FDR-q<0.25), including *Roseburia sp*., *Eubacterium sp*., *Lachnospira sp*., and *Coprobacter sp*.; but no bacterial genera were found to be associated with KYNA. Furthermore, an IPA-associated-bacteria score was inversely associated with plaque (OR=0.47[0.28, 0.79], *P*<0.01). But no significant effect modification by HIV serostatus was observed in these associations.

**Conclusions:** In a cohort of women living with and without HIV infection, plasma IPA levels and related gut bacteria were inversely associated with carotid artery plaque, suggesting a potential beneficial role of IPA and its gut bacterial producers in atherosclerosis and CVD.

## 1. Introduction

Tryptophan is an essential amino acid for internal protein synthesis and the biosynthesis of serotonin and melatonin in mammals. In humans, with the help of tryptophan 2,3-dioxygenase (TDO) and indoleamine 2,3-dioxygenase1 (IDO1), tryptophan is mainly catabolized through the kynurenine (KYN) pathway (1,2), in which several metabolites are produced, such as KYN and kynurenic acid (KYNA). Tryptophan can also be metabolized into a broad range of bioactive molecules, including indole and its derivatives [e.g., indole-3-lactic acid (ILA), indole-3-acid-acetic (IAA), and indole-3-propionic acid (IPA)], through the gut microbiota-dependent indole pathway (1,2). Some of these metabolites (e.g., IPA) exhibit antioxidative and anti-inflammatory properties (1,2), and therefore might have potential beneficial effects on human health.

Dysregulation of host tryptophan metabolism, particularly the depletion of tryptophan and enhancement of the KYN pathway, has been noted in human immunodeficiency virus (HIV) infection (3,4). Our work and other previous studies found that enhanced tryptophan catabolism, measured by KYN-to-tryptophan ratio (KYN/TRP) or KYNA-to-tryptophan ratio (KYNA/TRP), in people living with HIV (PLWH) was associated with subclinical atherosclerosis indicated by carotid artery plaque (5-7). These observations were in line with findings from studies of general populations (8-10), supporting the negative impacts of disrupted tryptophan metabolism and metabolites generated from the KYN pathway on cardiovascular disease (CVD) (11).

However, previous studies on tryptophan metabolism in relation to HIV infection and atherosclerosis were predominantly limited to examination of the KYN pathway metabolites (5,11,12); to date, whether other tryptophan metabolites (e.g., indole and the derivatives) are altered in PLWH, and their associations with atherosclerosis remains unclear. Furthermore, despite the well-known role of gut microbiota in tryptophan metabolism (1,2), no prior studies have integrated data on gut microbiota and tryptophan metabolites in relation to atherosclerosis among PLWH. Here, we aimed to investigate the interrelationship between tryptophan metabolites, gut microbiota, and carotid artery plaque among women living with and without HIV in the Women’s Interagency HIV Study (WIHS) (13).

## 2. Methods

### 2.1 Study design and population

The WIHS was a multicenter longitudinal study of women living with or at high risk for HIV infection designed to investigate the long-term, natural, and treated history of HIV infection (13), and now is a part of the Multicenter AIDS Cohort Study (MACS)/WIHS Combined Cohort Study (MWCCS) (14). Semi-annual visits consisted of structured questionnaire interviews, physical examinations, and specimen collection. In 2015, a multi-omics study was initiated among WIHS participants aged ≥ 35 years at three sites (Bronx, Brooklyn, and Chicago) to evaluate the etiology and progression of carotid artery atherosclerosis (15). Among them, 361 provided fecal samples for microbiome profiling and received assessments of carotid artery plaque, and 251 had plasma tryptophan metabolites measured. These participants were included in the present cross-sectional analyses. Participants who took antibiotics within the 4 weeks of stool sample collection were excluded from the microbiome association analyses. Detailed descriptions on the final sample sizes according to specific analyses are listed in the **Supplementary Fig 1**.

### 2.2 Assessment of HIV infection and other variables

Data on demographic, behavioral, medication, clinical, and laboratory biochemistry variables were collected following standardized protocols during the semi-annual visits. HIV infection was ascertained by enzyme-linked immunosorbent assay with confirmation using Western blot. HIV-specific parameters included CD4+ T-cell counts and HIV-1 viral load (measured by multiparametric flow cytometry of peripheral blood mononuclear cells) and use of antiretroviral therapy (ART). Cardiometabolic traits included body mass index (BMI), total cholesterol (TC), high-density lipoprotein (HDL) cholesterol, systolic blood pressure (SBP), fasting glucose, and use of lipid-lowering, antihypertensive, and antidiabetic medications.

### 2.3 Ascertainment of carotid artery plaque

High-resolution B-mode carotid artery ultrasound following a standardized protocol was used to image 8 locations in the right carotid artery: the near and far walls of the common carotid artery, carotid bifurcation, and internal and external carotid arteries (15,16). Focal plaque measures were obtained at a centralized reading center (University of Southern California). A focal plaque case was defined as an area with localized intima media thickness (IMT) > 1.5 mm in any of the 8 imaged carotid artery locations.

### 2.4 Measurement of tryptophan metabolites

Tryptophan metabolites were measured in blood plasma collected at the WIHS study visit closest to the date of carotid artery plaque assessment using a Hydrophilic Interaction Liquid Chromatography (HILIC)/Mass Spectrometry (MS) platform. Ten tryptophan metabolites were quantified, including tryptophan, KYNA, KYN, quinolinate, xanthurenate, serotonin, IPA, ILA, indoxylsulfate, and IAA. Raw data were processed using TraceFinder software (Thermo Fisher Scientific) for targeted peak integration and manual review of a subset of identified metabolites, which were then confirmed using authentic reference standards. In this study, all 10 metabolites were detected in more than 75% of participants. Levels of metabolites below detection were imputed by one half of the minimum detectable level of that metabolite and then were transformed by inverse-normal transformation (INVT) to improve normality and reduce the impacts of extreme values in association analyses. To assess the relative production of IPA, KYNA, and KYN from tryptophan, three ratios were constructed: IPA/TRP, KYNA/TRP, and KYN/TRP. In addition, the ratio of IPA to KYNA (IPA/KYNA) was calculated to indicate the switch of tryptophan metabolism from the production of KYNA to production of IPA.

### 2.5 Gut microbiome profiling

Fecal samples were collected using a home-based self-collection kit during WIHS core-visit as described previously (15,17). 16S ribosomal RNA (rRNA, V4 region) sequencing was performed on DNA extracted from fecal samples to profile the gut microbiome. After quality control, average coverage was approximately 35000 reads per sample. Details on the 16S rRNA sequencing method used in this study have been published (15,17). In addition, shotgun metagenomic sequencing based on a novel shallow-coverage method using the Illumina NovaSeq platform was performed in the same fecal samples among 141 women who had tryptophan metabolites data. De-multiplexing was applied to generate a shallow shotgun per-sample FASTQ dataset, and the adapter sequences were trimmed. Information on microbial taxonomic and functional profiles were generated using the shallow-shotgun computational (SHOGUN) pipeline (18).

### 2.6 Statistical analysis

Study population characteristics were described as means (standard deviations, SDs) or medians (inter-quartile ranges, IQRs) for continuous variables and frequencies (proportions) for categorical variables. Differences in characteristics between women with and without carotid artery plaque were compared using the Kruskal-Wallis test (for continuous variables) and Chi-squared test (for categorical variables). Logistic regression was used to assess the cross-sectional associations of tryptophan metabolites (inverse normal transformed and then scaled by dividing 1 SD) and the calculated ratios with prevalent carotid artery plaque, adjusting for age, race/ethnicity, education, smoking, study site and HIV serostatus (Model 1). We further adjusted for conventional cardiometabolic risk factors, including BMI, TC, HDL, SBP, and fasting glucose, and use of antihypertensive, lipid-lowering, and antidiabetic medications (Model 2). Stratified analyses were conducted to assess potential effect modification by HIV serostatus.

Analysis of Compositions of Microbiomes with Bias Correction (ANCOMBC) (19) was performed to identify microbiome genera associated with tryptophan metabolites with adjustment for the same set of covariates in the above Model 1. Herein, predominant genera presented in > 10% of samples with a 0.01% or higher relative abundance were included. Genera with a false discovery rate adjusted *P* value (FDR-q) < 0.25 were considered to be significantly differentiated ones. Shotgun metagenomic data were further leveraged to determine which species may drive the associations between identified genera and a given tryptophan metabolite in ANCOMBC. Multivariable linear regression was used to examine associations between bacterial species under the selected genera and tryptophan metabolites. In addition, we performed ANCOMBC to examine associations of KEGG Ortholog groups (KOs) with tryptophan metabolites. To assess joint associations of multiple genera associated with a given metabolite with carotid artery plaque, a weighted bacteria score was constructed as the sum of the products of central log ratio (CLR)-transformed abundances of identified genera and their corresponding coefficients in ANCOMBC. We then examined the cross-sectional associations of selected individual genera and the bacterial score with carotid artery plaque using logistic regression, adjusting for covariates listed in the above Model 1. These analyses were limited to tryptophan metabolites that were associated with both microbiome genera and carotid artery plaque.

All analyses were conducted in R 3.4. Statistical significance was set at 0.05 (two-tailed) unless otherwise stated.

## 3. Results

### 3.1 Study population characteristics

Among women with both carotid artery plaque and 16S rRNA sequencing data (n=361), 241 were living with HIV (67%) and 98 (27%) had carotid artery plaque (**Table 1**). Compared with those without plaque, women with plaque were older (median: 58 vs 52 yrs) and were more likely to take lipid-lowering (34% vs 20%), antihypertensive (60% vs 42%) and antidiabetic (27.6% vs 15.6%) medications. No significant differences in other characteristics, including HIV-related variables, were found between these two groups. Similar patterns were observed when comparisons were limited to participants with collective data on tryptophan metabolites and plaque (n=188) (**Supplementary Table 1**).

**Table 1.** Population characteristics by carotid artery plaque status (N=361)

| Characteristics | Without plaque<br>(n = 263) | With plaque<br>(n = 98) | *P*<sup>a</sup> |
| --- | --- | --- | --- |
| Age (years) | 51.6 (7.9) | 57.5 (7.0) | <0.001 |
| BMI (kg/m<sup>2</sup>) | 31.9 (7.9) | 30.9 (7.8) | 0.18 |
| Race (n,%) |  |  | 0.95 |
| African American | 187 (71.1%) | 68 (69.4%) |  |
| Hispanic | 59 (22.4%) | 23 (23.5%) |  |
| White/others | 17 (6.5%) | 7 (7.1%) |  |
| Education levels (n, %) |  |  | 0.61 |
| Before Highschool | 121 (46%) | 43 (43.9%) |  |
| Highschool | 80 (30.4%) | 27 (27.6%) |  |
| College and above | 62 (23.6%) | 28 (28.6%) |  |
| Family annual income (n, %) |  |  | 0.76 |
| <12,000\$ | 158 (60%) | 63 (64.3%) | |
| 12,000~24,000\$ | 59 (22.4%) | 20 (20.4%) | |
| >24,000\$ | 46 (17.5%) | 15 (15.3%) | |
| Smoking status (n,%) |  |  | 0.08 |
| Never smoker | 59 (22.4%) | 14 (14.3%) |  |
| Current smoker | 109 (41.4%) | 37 (37.8%) |  |
| Former smoker | 95 (36.1%) | 47 (47.9%) |  |
| Total cholesterol (mg/dL) | 187.9 (36.6) | 188.9 (36.6) | 0.78 |
| HDL cholesterol (mg/dL) | 57.6 (17.9) | 57.7 (19.0) | 0.98 |
| Systolic blood pressure (mmHg) | 127.8 (19.4) | 134.4 (21.9) | 0.01 |
| Fasting glucose (mg/dL) | 98.0 (26.7) | 100.7 (33.1) | 0.84 |
| Took lipids lowering medication<br>(n,%) | 52 (19.8%) | 33 (33.7%) | 0.06 |
| Took antihypertensive medication<br>(n, %) | 111 (42.2%) | 59 (60.2%) | 0.02 |
| Took antidiabetic medication (n,%) | 41 (15.6%) | 27 (27.6%) | 0.01 |
| HIV positive (n,%) | 172 (65.4%) | 69 (70.4%) | 0.40 |
| Highly active ART use in past 6<br>months among HIV+<sup>b</sup> | 157 (91.2%) | 65 (94.2%) | 0.40 |
| CD4 positive cells (per mm<sup>3</sup>)<sup>b</sup> | 679.0 (429.3, 923.8) | 646.0 (523.0, 860.0) | 0.90 |
| Detectable HIV RNA (n,%)<sup>b</sup> | 52 (30.4%) | 18 (26.1%) | 0.61 |
| HIV RNA virus load (cp/mL)<sup>c</sup> | 71.0 (36.5, 1,912.5) | 43.0 (33.50, 224.0) | 0.40 |
| Antibiotics use (n,%)<sup>d</sup> | 12 (4.6%) | 4 (4.1%) | 0.98 |

Notes: <sup>a</sup> *P* values were generated according to student Student's t-test (for normally distributed variables), Kruskal–Wallis test for (for highly skewed variables), and Chi-squared Test (for categorical variables); <sup>b</sup> only available among HIV+; <sup>c</sup> only among participants with HIV RNA detectable. Continuous variables were presented as mean (standard deviation [SD]), except for CD4 positive cells and HIV RNA virus load, which were presented as median (interquartile range, IQR) due to their highly skewed distributions; <sup>d</sup> participants with antibiotic use were excluded in the microbiome analyses.

The Spearman correlation matrix among tryptophan metabolites (**Supplementary Fig 2**) revealed moderate correlations (0.11<r_*s*_<0.54) among those produced by KYN pathway (KYN, quinolinate, KYNA, and xanthurenate), but relatively weak correlations among indole derivatives (−0.05<r_*s*_<0.23). Plasma KYN, quinolinate, and indoxysulfate levels as well as KYN/TRP and KYNA/TRP were higher among women with HIV as compared to those without HIV; but no differences were found for other metabolites and ratios by HIV serostatus (**Supplementary Fig 3**).

### 3.2 Associations between tryptophan metabolites and carotid artery plaque

Among 10 tryptophan metabolites, only KYNA (OR=1.93, 95%CI: 1.12, 3.32 per one SD increment) and IPA (OR=0.62, 95%CI: 0.40, 0.98) were associated with carotid artery plaque in Model1 (**Fig 1**). We also found that KYNA/TRP was positively associated with plaque (OR=1.83, 95%CI: 1.08, 3.09), while IPA/ KYNA presented inverse associations with plaque (OR=0.51, 95%CI: 0.33, 0.80). The association between IPA and plaque was attenuated and no longer significant, while other significant associations remained after further adjusting for conventional cardiometabolic risk factors (**Fig 1**). Further analysis indicated that the attenuation on the association of IPA might be due to the further adjustment for diabetes related factors (e.g., fasting glucose and antidiabetic medications use) (**Supplementary Table 2**).

**Fig 1.**
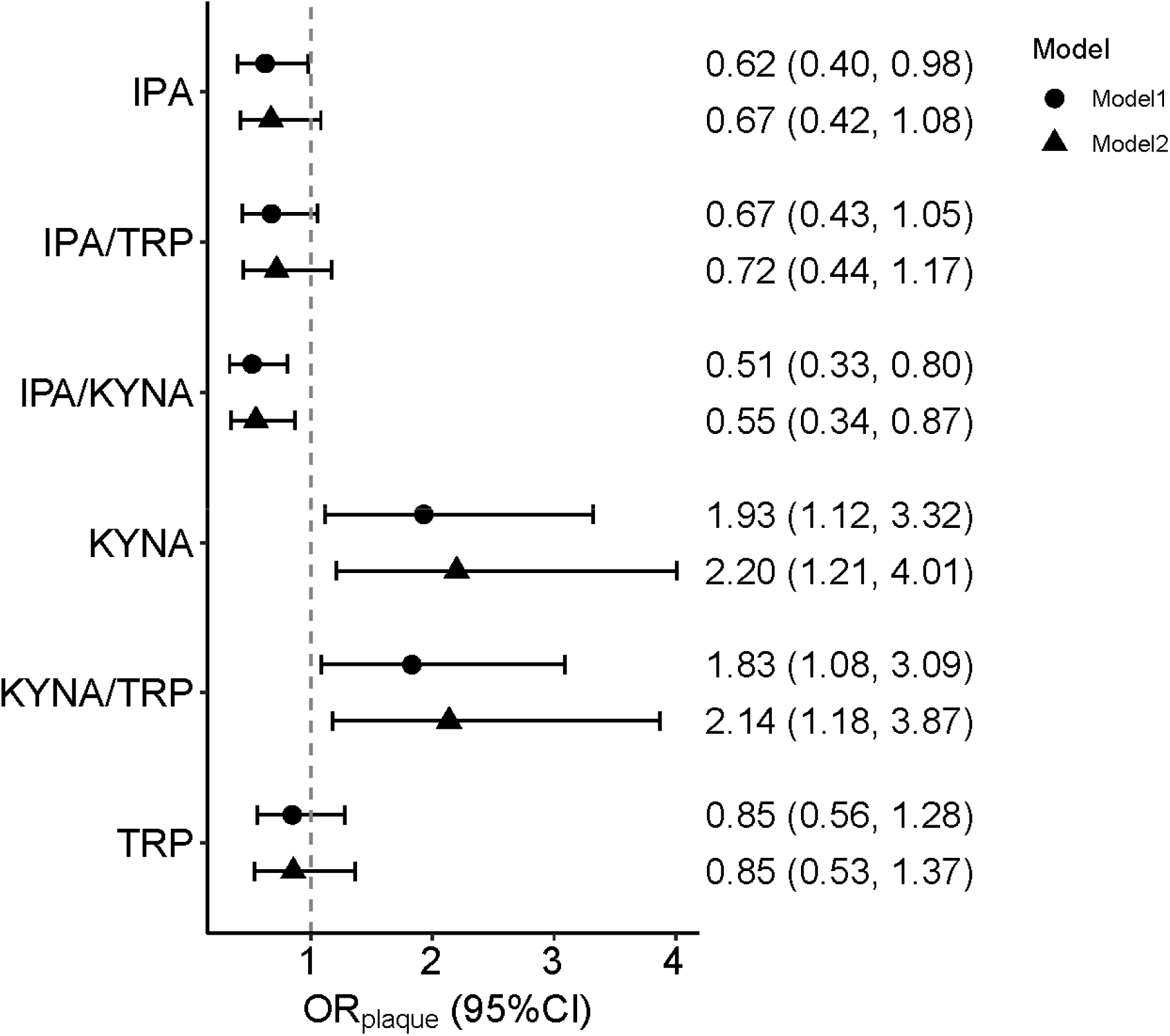
Associations of selected tryptophan (TRP) metabolites and derived ratios with prevalent carotid artery plaque (N=188). Estimates [odds ratio of carotid artery plaque (OR_plaque_) and 95% confidence interval (CI)] per one SD increasement in TRP metabolites concentrations (after inverse normal transformation) based on logistic regression. Model 1 was adjusted for age, race/ethnicity, study sites, education, smoking and HIV serostatus at visit. Model 2 was additionally adjusted for BMI, total cholesterol, HDL cholesterol, systolic blood pressure, fasting glucose, took antihypertensive medication, took lipids lowering medication, and took antidiabetic medication. Results of the other TRP metabolites were presented in the Supplementary Table 2. Abbreviations: IPA: indole-3-propionate; TRP: tryptophan; KYNA: kynurenic acid. IPA/TRP, KYNA/TRP, and IPA/KYNA were the ratios of the corresponding two metabolites.

Stratified analysis by HIV serostatus revealed that the associations of IPA and two ratios of IPA (IPA/TRP and IPA/KYNA) with carotid artery plaque tended to be stronger among women living with HIV compared to those without HIV, though no significant effect modification was found (**Supplementary Table 3**). For example, IPA was significantly associated with carotid artery plaque in women living with HIV (OR=0.40, 95%CI: 0.20, 0.80) but not in those without HIV (OR=0.88, 95%CI: 0.39, 1.99) (*P*-_for interaction_=0.078).

### 3.3 Associations between gut microbiota features and tryptophan metabolites

Using 16s rRNA sequencing data among 205 participants without use of antibiotics, we identified 20 bacterial genera (FDR-q<0.25) associated with 7 tryptophan metabolites (**Supplementary Table 4**). Among them, 6 genera were associated with IPA, with positive associations being observed for 2 genera in the *Firmicute* phylum (i.e., *Roseburia* and *Lachnospira*) and three unclassified groups (*Barnesiellaceae_unclassified, Clostridiales_unclassified*, and *RF39_unclassified*), and an inverse association for *Eggerthella* (**Fig 2A**). The other 14 genera were associated with tryptophan (n=5), IAA (n=4), indoxylsufate (n=3), xanturenate (n=2), KYN (n=1), and quinolinate (n=1) (**Supplementary Table 4**). Most of these bacterial genera were associated with only one metabolite (**Fig 2B** and **Supplementary Table 4**).

**Fig 2.**
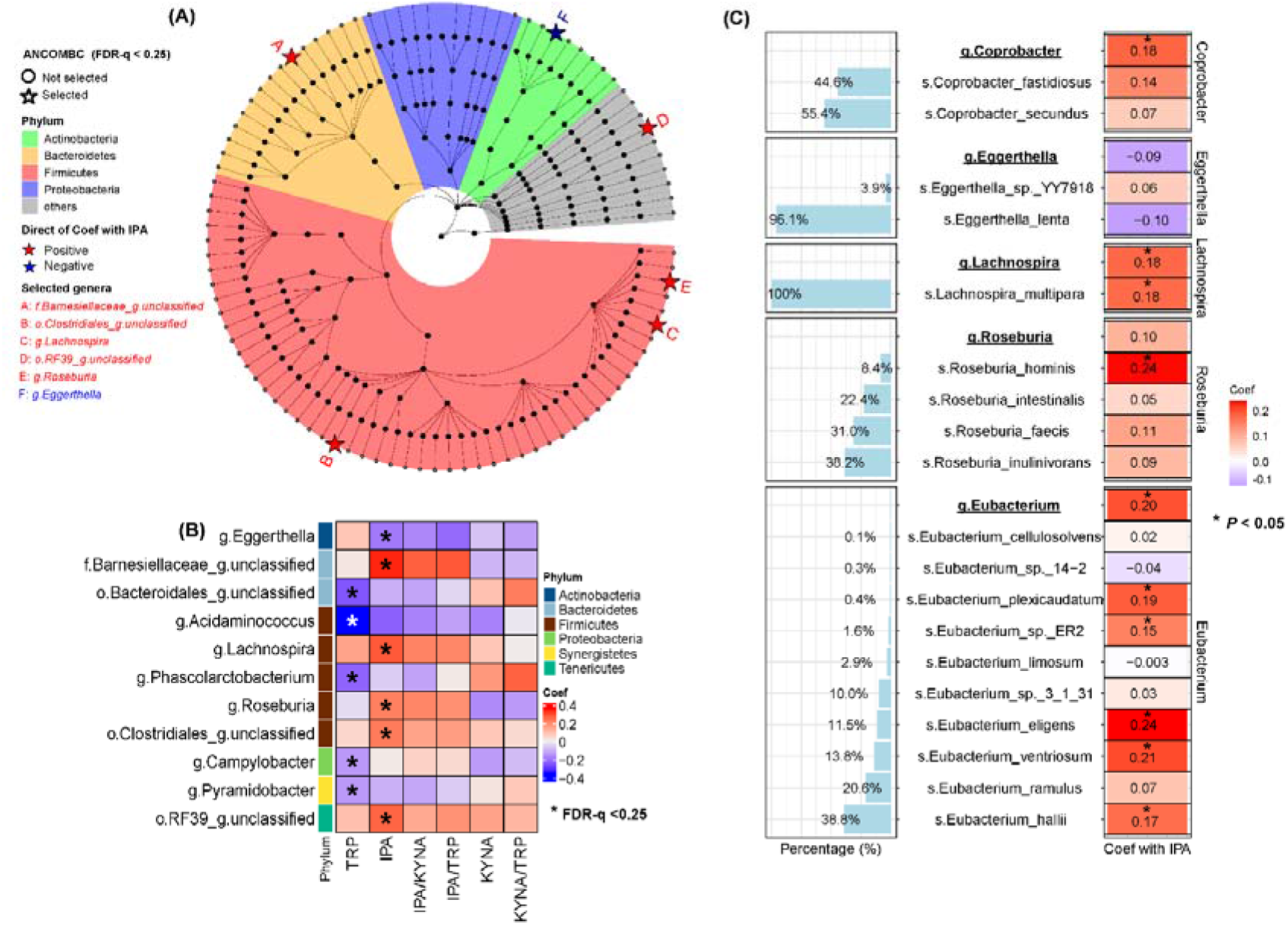
Taxonomic features associated with selected plasma tryptophan (TRP) metabolites. (A) presents the phylogenetic tree for taxonomic features of the included 97 predominant genera in the ANCOMBC analysis for associations with tryptophan metabolites (n=205). Stars with letters (from A to F) refer to indole-3-propionate (IPA) -associated genera (FDR-q<0.25). (B) presents the heatmap of the coefficients of associations of IPA or tryptophan associated genera with tryptophan, IPA, kynurenic acid (KYNA), IPA/TRP, IPA/KYNA, and KYNA/TRP in ANCOMBC while adjusting for age, race/ethnicity, study sites, education, smoking, and HIV serostatus at visit. The numeric results were presented in the Supplementary Table 4. (C) presents the relationship (coefficients per SD increment in taxa) between the identified species within IPA-associated genera (bolded and underlined) and IPA levels among participants with shotgun sequencing data (n=136) (the right heatmap). The left boxplot presents the percentages of these species within the specific genus. Prefixes of “o.”, “f.”, “g.”, and “s.” refer to the taxa at the order, family, genus and species levels, respectively. The numeric results were presented in the Supplementary Table 5.

We then focused on IPA-associated bacterial genera given that IPA was the only microbiota-dependent metabolite that was associated with carotid artery plaque in this study. We used metagenomic sequencing data in a subsample (n=136) to identify which bacterial species might drive the observed IPA-genera associations. To characterize three unclassified genera, we first examined the genera under the *Barnesiellaceae* family and those under the *Clostridiales* and *RF39* order that were not captured in the 16S rRNA data. Herein, we additionally identified *Corprobacter* genus under the *Barnesiellaceae* family (**Fig 2C**), and 7 genera of 4 families under the *Clostridiales* order (**Supplementary Fig 4**) in the metagenomics data. We found that the associations of *Barnesiellaceae_unclassified* with IPA tended to be driven by *Coprobacter* genus (*P*<0.05), possibly by *Coprobacter_fastidiosus* species (**Fig 2C**). Associations of *Clostridiale_unclassified* tended to be driven by *Eubacterium* genus (**Supplementary Fig 4**) and affiliated species (**Fig 2C**) within the *Eubacteriaceae* family, such as *Eubacterium_eligens* and *Eubacterium_ventriosum*. But no bacteria within the *RF39* order were identified in metagenomic data. For genera *Roseburia, Lachnospira* and *Eggerthella*, their associations with IPA tended to be driven by *Roseburia_hominis, Lachnospira_multipara* and *Eggerthella_lenta*, respectively (**Fig 2C** and **Supplementary Table 5**). In line with the results on plasma IPA levels between women living with and without HIV infection, no significant differences were found for abundances of the 6 IPA-associated genera or species by HIV serostatus (data not shown).

Further analysis of metagenomic function data identified 189 KOs (FDR-q<0.05) associated with IPA (**Supplementary Fig 5A**), but none of them seem to be involved in the metabolic pathway from tryptophan to IPA when searching the MetaCyc database. We found two enzymes (i.e., Cs-FLDA (3-(aryl) acryloyl-CoA:(R)-3-(aryl) lactate CoA-transferase) [K13607] and ACDS (butyryl-CoA dehydrogenase) [K00248]) that are involved in this pathway in our metagenomics data, but they were not associated with IPA levels (**Supplementary Fig 5B**).

### 3.4 Associations between indole-3-propionate associated bacteria and carotid artery plaque

We further linked the 6 IPA-associated bacteria genera with plaque and found that genera having positive associations with IPA (**Fig 3A**) largely tended to be inversely associated with plaque (**Fig 3B**). By constructing a weighted genera score with the inclusion of the identified 6 genera and the other two previously reported IPA-associated genera (i.e., *Adlercreutzia* and *Faecalibacterium*) (20) that were marginally associated with IPA in the current study (**Supplementary Table 4**), we found that the constructed IPA-associated bacteria score was significantly positively associated with both plasma IPA levels and IPA/KYNA (**Fig 3A**) and was inversely associated with plaque (**Fig 3B**). This association with plaque was attenuated when further adjusting for IPA or IPA/KYNA (**Fig 3C and 3D**), suggesting that the association might be partly explained by plasma IPA and enhanced IPA production. Similar association patterns between IPA-associated bacteria score and plaque were observed among 345 women with both plaque and 16S rRNA sequencing data (**Supplementary Table 6**).

**Fig 3.**
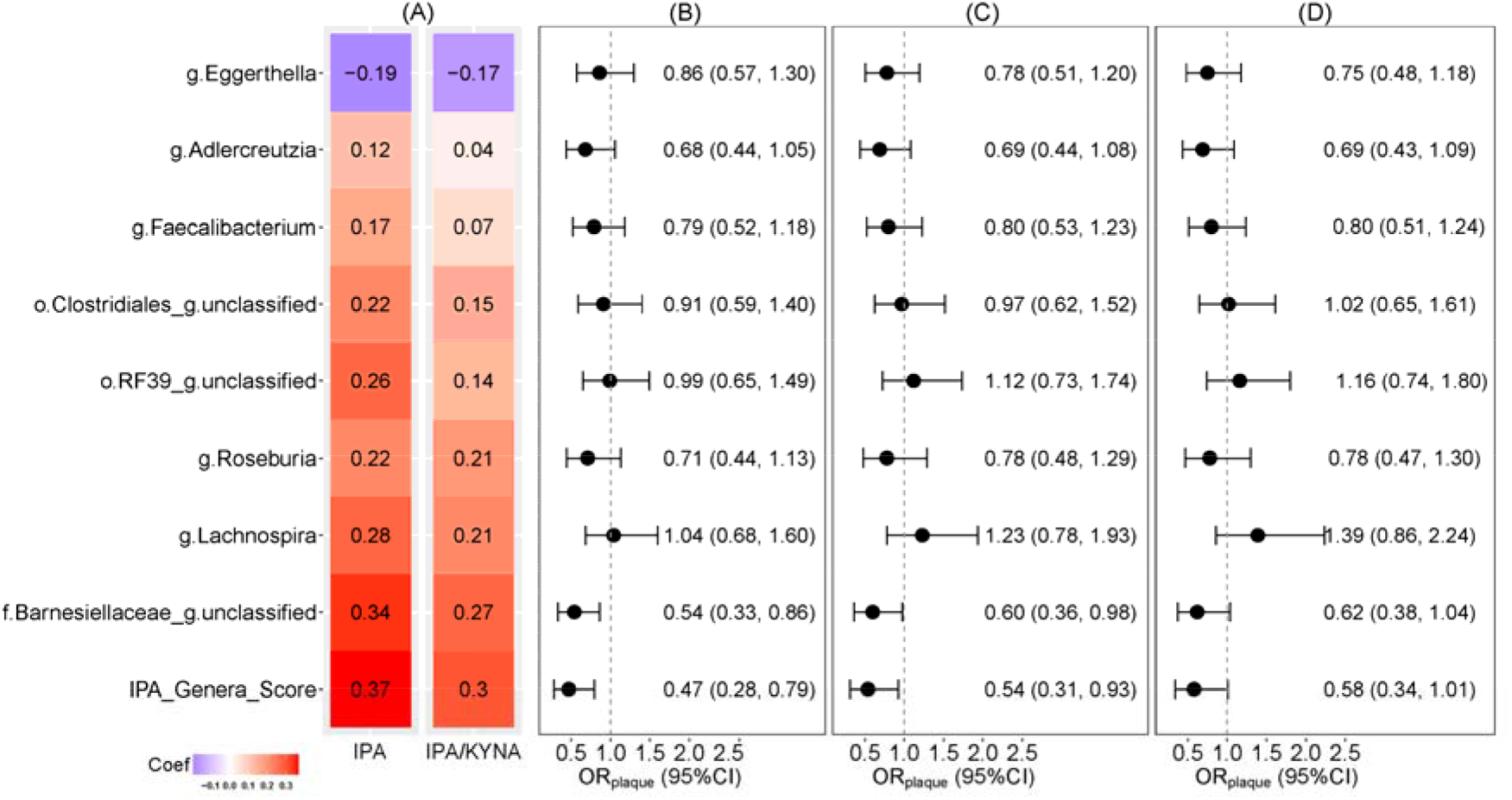
Associations of indole-3-propionate (IPA) associated-genera with prevalent carotid artery plaque. OR_plaque_ refer to the odds ratio (OR) of carotid artery plaque. (A) presents the regression coefficients for the associations of selected IPA-associated genera and IPA_genera_score with IPA levels and IPA-to-kynurenic acid ratio (IPA/KYNA) (n=205). (B) presents the associations of IPA associated genera and IPA_genera_score among participants with collective data on carotid artery plaque, 16S rRNA sequencing and IPA levels (n=163, no of plaque cases =38). (C) associations additionally adjusted for IPA levels. (D) associations additionally adjusted for IPA/KYNA. Estimates in (A) to (D) were all adjusted for age, race/ethnicity, study sites, education status, smoking status, and HIV serostatus at visit. Abundances of genera were centered log ratio (CLR) transformed and then were scaled into Z-scores; the IPA_genera_score were the weighted sum of CLR values of selected genera, where the weights were their corresponding coefficients in ANCOMBC analysis. The weights of genera that were negatively associated with IPA concentrations were reversed in the calculation of IPA_genera_score. *Adlercreutzia* and *Faecalibacterium* are the two previously reported IPA-associated genera (doi:10.1136/gutjnl-2021-324053) but were marginally associated with IPA levels in present analysis (*P* = 0.067 and *P* = 0.056, respectively). Prefixes of “o.”, “f.”, and “g” refer to the taxa at the order, family, and genus levels, respectively.

## 4. Discussion

In this study of women with or at risk for HIV infection, we found that plasma KYNA and KYNA/TRP were positively associated with carotid artery plaque, independent of HIV serostatus and traditional cardiovascular risk factors, which is line with our previous among MWCCS participants during earlier visits (5). Furthermore, we found that plasma IPA and IPA/KYNA were inversely associated with carotid artery plaque, with suggestive stronger associations among women living with HIV compared to those without HIV. We identified multiple gut bacteria associated with plasma IPA and the IPA-associated bacteria score were also inversely associated with plaque.

Epidemiological studies either in HIV or non-HIV populations have linked enhanced KYN pathway of tryptophan metabolism with multiple atherosclerotic effects, such as increased macrophage-loaded cores of atherosclerotic plaques (21), carotid artery intima-media thickness (IMT) (6), and elevated risk of myocardial infarction (11). Together with these observations and our previous findings (5), our results highlight the potential of metabolites (e.g., KYNA and KYN) generated from the KYN pathway associated with elevated risk of CVD. Although gut microbiota might be involved in the tryptophan-KYN metabolism pathway in the gut (1,12), we did not find many associations between gut bacteria and plasma KYN pathway metabolites. Unlike indole metabolites (e.g., IPA) which are microbiota dependent, circulating KYN pathway metabolites might be mainly attributed to host tryptophan metabolism (1). However, this could also be partly due to limitations of our 16S rRNA sequencing data that might not capture gut bacteria related to the production of KYN metabolites.

The observed protective association between IPA and carotid artery plaque in our study is consistent with current evidence in the literature that IPA is generally beneficial to the cardiovascular system, possibly through suppression of proinflammatory cytokine expression (22-24), promotion of macrophage reverse cholesterol transport (25), and reduction of cellular reactive oxygen species (26,27), oxidative damage and lipid peroxidation (28,29).

Our results are also supported by previous findings in studies among general populations, wherein lower plasma IPA levels were observed in patients with advanced atherosclerosis as compared to healthy controls (25,30). Several prospective studies have linked higher circulating IPA levels with lower risk of diabetes (20,31,32). In line with this, we also found that the association between IPA and plaque could be partially explained by traditional CVD risk factors, especially those related to diabetes. Interestingly, we observed a stronger association between IPA and plaque among women living with HIV compared to those without HIV; however, the statistical interaction test was borderline significant, which might be due to the limited sample size. This result suggests that IPA might have extra favorable relationship with cardiovascular health in people living with HIV, but further studies are warranted to validate our findings and examine the underlying mechanisms.

Several bacteria, mostly in the Firmicutes phylum, have been found to be involved in the production of IPA (2,33). Our study identified multiple bacterial genera, including those previously reported (e.g., *Roseburia, Eubacterium, Faecalibacterium, Lachnospira*, and unknown genera under the *RF39* order) (2,20,34) and potentially novel ones (e.g., *Coprobacter*) associated with circulating IPA levels. Consistently, most of these bacteria belongs to Firmicutes, and their positive associations with IPA might be related to host dietary fiber intake (20). *Roseburia, Eubacterium, Faecalibacterium* and *Lachnospira* are known for fiber degradation (35,36) and *Coprobacter* can be increased following laminaran supplementation in rats (37). High fiber intake thus could increase the population of these potential IPA producers in the gut, which in turn could shift tryptophan metabolism toward IPA production (20). However, the lack of dietary data in our study prevents further testing of this hypothesis. In addition, although several bacterial enzymes have been suggested to be involved in the IPA production pathway (33), only two KOs of these enzymes were detected in our metagenomic data, with relatively small abundance. They were not associated with plasma IPA, which might be due to gaps between the measured abundances of bacterial genes encoding enzymes and the real enzyme activity in the bacterial metabolite production (38). Regarding the role of IPA-associated bacteria in the progression of atherosclerosis, other potential mechanisms beyond IPA production could also be possible since the association between IPA-associated bacteria score and carotid artery plaque were not fully explained by IPA levels. For examples, many of these fiber-utilizing bacteria (e.g., *Roseburia, Eubacterium, Faecalibacterium* and *Lachnospira)* are also producers of shot-chain fatty acids that might be beatifical to cardiovascular health (39).

Major strengths of this study include the relatively broad assessment of tryptophan metabolites, including both KYN pathway metabolites and indole derivatives, and the integrative analysis of gut microbiota with tryptophan metabolites in relation to carotid artery plaque. Nevertheless, several caveats should be acknowledged. First, the moderate sample size prevented us from conducting comprehensive stratified analyses to assess potential effect modification by HIV serostatus and other interesting HIV infection related characteristics. Second, the gut microbiome profiling predominantly relied on the 16S rRNA sequencing, which has a relatively low taxonomy resolution and insufficient information on microbial functional profiles (40). Although our shallow shotgun metagenomics in a subsample revealed interesting data at species level, it was still limited to examine microbial functional components. Third, the lack of dietary data further hinders us from assessing the microbiota-diet interaction in the relationship between tryptophan metabolites and carotid artery plaque. Finally, the nature of this observational investigation with a cross-sectional design precludes causal inference regarding the observed associations.

## 5. Conclusions

This cross-sectional study among women from the WIHS extended our prior findings on the unfavorable associations between tryptophan-KYN pathway metabolites and carotid artery atherosclerosis (5) to demonstrate a favorable relationship between IPA, a microbially produced metabolite from tryptophan, and carotid artery atherosclerosis, especially in women with HIV infection. Gut bacteria associated with higher IPA levels were also favorably linked with carotid artery plaque. These findings suggest a potential beneficial role of IPA and associated gut microbiota in atherosclerosis and CVD. Further prospective studies with larger sample sizes and metagenomics data are warranted to confirm our findings and reveal underlying mechanisms.

## Supporting information

Supplemental figures and description

## Data Availability

Any requests as to the potential replication or source data should be addressed to the corresponding author.

## Abbreviations

HIV: human immunodeficiency virus
WIHS: Women’s Interagency HIV Study
MACS: Multicenter AIDS Cohort Study
(MWCCS): MACS-WIHS Combined Cohort Study
CVD: cardiovascular disease
FDR: false discovery rate
TRP: tryptophan
KYN: kynurenine
KYNA: kynurenic acid
ILA: indole-3-lactic acid
IAA: indole-3-acid-acetic
IPA: indole-3-propionic acid
HILIC: Hydrophilic Interaction Liquid Chromatography
MS: Mass Spectrometry
SHOGUN: shallow-shotgun
IQR: inter-quartile range
SD: standard deviation
ANCOMBC: Analysis of Compositions of Microbiomes with Bias Correction
(KOs): KEGG Ortholog groups
CLR: central log ratio
Cs-FLDA: 3-(aryl) acryloyl-CoA:(R)-3-(aryl) lactate CoA-transferase
ACDS: butyryl-CoA dehydrogenase
OR: odds ratio
ART: antiretroviral therapy
BMI: body mass index
TC: total cholesterol
HDL: high-density lipoprotein
SBP: systolic blood pressure

## Supplementary Information

Supplementary Tables and Figs can be found at online.

## CRediT authorship contribution statement

Kai Luo: Conceptualization, Formal analysis, Methodology, Writing – original draft. Zheng Wang, Tao Wang, Christopher C Sollecito, Evan Grassi, Fanua Wiek, Lauren St Peter, Mykhaylo Usyk, and Clary B Clish: Methodology and data curation. Brandilyn A Peters, David B Hanna, Wendy S Post, Alan L Landay, Howard N Hodis, Kathleen M Weber, Audrey French, Elizabeth T Golub, Jason Lazar, Deborah Gustafson, Anjali Sharma, Kathryn Anastos, Rob Knight, Robert C Kaplan and Robert D Burk: Writing – review & editing. Qibin Qi: Conceptualization, Supervision, Project administration, Funding acquisition, Writing – review & editing. The authors read and approved the final manuscript.

## Acknowledgments

Data in this manuscript were collected by the Women’s Interagency HIV Study (WIHS), now the MACS/WIHS Combined Cohort Study (MWCCS). The authors gratefully acknowledge the contributions of the study participants and dedication of the staff at the MWCCS sites.

## Disclaimer

The contents of this article are solely the responsibility of the authors and do not represent the official views of the National Institutes of Health (NIH).

## Ethnics declaration

All participants provided written informed content and the present study was reviewed and approved by the Institutional Review Board (IRB) at Albert Einstein College of Medicine (IRB no: 2018-9386).

## References

1. Agus A, Planchais J, Sokol H. Gut Microbiota Regulation of Tryptophan Metabolism in Health and Disease. Cell Host Microbe. 2018;23(6):716–724.

2. Roager HM, Licht TR. Microbial tryptophan catabolites in health and disease. Nat Commun. 2018;9(1):3294.

3. Gelpi M, Hartling HJ, Ueland PM, Ullum H, Troseid M, Nielsen SD. Tryptophan catabolism and immune activation in primary and chronic HIV infection. BMC Infect Dis. 2017;17(1):349.

4. Murray MF. Tryptophan depletion and HIV Tryptophan depletion and HIV infection: a metabolic link to pathogenesis. The Lancet infectious diseases. 2003;3(10):644–652.

5. Qi Q, Hua S, Clish CB, Scott JM, Hanna DB, Wang T, Haberlen SA, Shah SJ, Glesby MJ, Lazar JM, Burk RD, Hodis HN, Landay AL, Post WS, Anastos K, Kaplan RC. Plasma Tryptophan-Kynurenine Metabolites Are Altered in Human Immunodeficiency Virus Infection and Associated With Progression of Carotid Artery Atherosclerosis. Clin Infect Dis. 2018;67(2):235–242.

6. Boyd A, Boccara F, Meynard JL, Ichou F, Bastard JP, Fellahi S, Samri A, Sauce D, Haddour N, Autran B, Cohen A, Girard PM, Capeau J, Collaboration in Hiv I, Cardiovascular Disease s. Serum Tryptophan-Derived Quinolinate and Indole-3-Acetate Are Associated With Carotid Intima-Media Thickness and its Evolution in HIV-Infected Treated Adults. Open Forum Infect Dis. 2019;6(12):ofz516.

7. Siedner MJ, Kim JH, Nakku RS, Bibangambah P, Hemphill L, Triant VA, Haberer JE, Martin JN, Mocello AR, Boum Y, 2nd, Kwon DS, Tracy RP, Burdo T, Huang Y, Cao H, Okello S, Bangsberg DR, Hunt PW. Persistent Immune Activation and Carotid Atherosclerosis in HIV-Infected Ugandans Receiving Antiretroviral Therapy. J Infect Dis. 2016;213(3):370–378.

8. Kato A, Suzuki Y, Suda T, Suzuki M, Fujie M, Takita T, Furuhashi M, Maruyama Y, Chida K, Hishida A. Relationship between an increased serum kynurenine/tryptophan ratio and atherosclerotic parameters in hemodialysis patients. Hemodial Int. 2010;14(4):418–424.

9. Pawlak K, Brzosko S, Mysliwiec M, Pawlak D. Kynurenine, quinolinic acid--the new factors linked to carotid atherosclerosis in patients with end-stage renal disease. Atherosclerosis. 2009;204(2):561–566.

10. Sulo G, Vollset SE, Nygard O, Midttun O, Ueland PM, Eussen SJ, Pedersen ER, Tell GS. Neopterin and kynurenine-tryptophan ratio as predictors of coronary events in older adults, the Hordaland Health Study. Int J Cardiol. 2013;168(2):1435–1440.

11. Pedersen ER, Tuseth N, Eussen SJ, Ueland PM, Strand E, Svingen GF, Midttun O, Meyer K, Mellgren G, Ulvik A, Nordrehaug JE, Nilsen DW, Nygard O. Associations of plasma kynurenines with risk of acute myocardial infarction in patients with stable angina pectoris. Arterioscler Thromb Vasc Biol. 2015;35(2):455–462.

12. Vujkovic-Cvijin I, Dunham RM, Iwai S, Maher MC, Albright RG, Broadhurst MJ, Hernandez RD, Lederman MM, Huang Y, Somsouk M. Dysbiosis of the gut microbiota is associated with HIV disease progression and tryptophan catabolism. Science translational medicine. 2013;5(193):193ra191–193ra191.

13. Adimora AA, Ramirez C, Benning L, Greenblatt RM, Kempf MC, Tien PC, Kassaye SG, Anastos K, Cohen M, Minkoff H, Wingood G, Ofotokun I, Fischl MA, Gange S. Cohort Profile: The Women’s Interagency HIV Study (WIHS). Int J Epidemiol. 2018;47(2):393–394i.

14. D’Souza G, Bhondoekhan F, Benning L, Margolick JB, Adedimeji AA, Adimora AA, Alcaide ML, Cohen MH, Detels R, Friedman MR, Holman S, Konkle-Parker DJ, Merenstein D, Ofotokun I, Palella F, Altekruse S, Brown TT, Tien PC. Characteristics of the MACS/WIHS Combined Cohort Study: Opportunities for Research on Aging With HIV in the Longest US Observational Study of HIV. Am J Epidemiol. 2021;190(8):1457–1475.

15. Wang Z, Peters BA, Usyk M, Xing J, Hanna DB, Wang T, Post WS, Landay AL, Hodis HN, Weber K, French A, Golub ET, Lazar J, Gustafson D, Kassaye S, Aouizerat B, Haberlen S, Malvestutto C, Budoff M, Wolinsky SM, Sharma A, Anastos K, Clish CB, Kaplan RC, Burk RD, Qi Q. Gut Microbiota, Plasma Metabolomic Profiles, and Carotid Artery Atherosclerosis in HIV Infection. Arterioscler Thromb Vasc Biol. 2022;42(8):1081–1093.

16. Hanna DB, Post WS, Deal JA, Hodis HN, Jacobson LP, Mack WJ, Anastos K, Gange SJ, Landay AL, Lazar JM, Palella FJ, Tien PC, Witt MD, Xue X, Young MA, Kaplan RC, Kingsley LA. HIV Infection Is Associated With Progression of Subclinical Carotid Atherosclerosis. Clin Infect Dis. 2015;61(4):640–650.

17. Moon JY, Zolnik CP, Wang Z, Qiu Y, Usyk M, Wang T, Kizer JR, Landay AL, Kurland IJ, Anastos K, Kaplan RC, Burk RD, Qi Q. Gut microbiota and plasma metabolites associated with diabetes in women with, or at high risk for, HIV infection. EBioMedicine. 2018;37:392–400.

18. Hillmann B, Al-Ghalith GA, Shields-Cutler RR, Zhu Q, Gohl DM, Beckman KB, Knight R, Knights D. Evaluating the Information Content of Shallow Shotgun Metagenomics. mSystems. 2018;3(6).

19. Lin H, Peddada SD. Analysis of compositions of microbiomes with bias correction. Nat Commun. 2020;11(1):3514.

20. Qi Q, Li J, Yu B, Moon JY, Chai JC, Merino J, Hu J, Ruiz-Canela M, Rebholz C, Wang Z, Usyk M, Chen GC, Porneala BC, Wang W, Nguyen NQ, Feofanova EV, Grove ML, Wang TJ, Gerszten RE, Dupuis J, Salas-Salvado J, Bao W, Perkins DL, Daviglus ML, Thyagarajan B, Cai J, Wang T, Manson JE, Martinez-Gonzalez MA, Selvin E, Rexrode KM, Clish CB, Hu FB, Meigs JB, Knight R, Burk RD, Boerwinkle E, Kaplan RC. Host and gut microbial tryptophan metabolism and type 2 diabetes: an integrative analysis of host genetics, diet, gut microbiome and circulating metabolites in cohort studies. Gut. 2022;71(6):1095–1105.

21. Niinisalo P, Oksala N, Levula M, Pelto-Huikko M, Jarvinen O, Salenius JP, Kytomaki L, Soini JT, Kahonen M, Laaksonen R, Hurme M, Lehtimaki T. Activation of indoleamine 2,3-dioxygenase-induced tryptophan degradation in advanced atherosclerotic plaques: Tampere vascular study. Ann Med. 2010;42(1):55–63.

22. Garcez ML, Tan VX, Heng B, Guillemin GJ. Sodium Butyrate and Indole-3-propionic Acid Prevent the Increase of Cytokines and Kynurenine Levels in LPS-induced Human Primary Astrocytes. Int J Tryptophan Res. 2020;13:1178646920978404.

23. Zhao ZH, Xin FZ, Xue Y, Hu Z, Han Y, Ma F, Zhou D, Liu XL, Cui A, Liu Z, Liu Y, Gao J, Pan Q, Li Y, Fan JG. Indole-3-propionic acid inhibits gut dysbiosis and endotoxin leakage to attenuate steatohepatitis in rats. Exp Mol Med. 2019;51(9):1–14.

24. Venkatesh M, Mukherjee S, Wang H, Li H, Sun K, Benechet AP, Qiu Z, Maher L, Redinbo MR, Phillips RS, Fleet JC, Kortagere S, Mukherjee P, Fasano A, Le Ven J, Nicholson JK, Dumas ME, Khanna KM, Mani S. Symbiotic bacterial metabolites regulate gastrointestinal barrier function via the xenobiotic sensor PXR and Toll-like receptor 4. Immunity. 2014;41(2):296–310.

25. Xue H, Chen X, Yu C, Deng Y, Zhang Y, Chen S, Chen X, Chen K, Yang Y, Ling W. Gut Microbially Produced Indole-3-Propionic Acid Inhibits Atherosclerosis by Promoting Reverse Cholesterol Transport and Its Deficiency Is Causally Related to Atherosclerotic Cardiovascular Disease. Circ Res. 2022:101161CIRCRESAHA122321253.

26. Poeggeler B, Pappolla MA, Hardeland R, Rassoulpour A, Hodgkins PS, Guidetti P, Schwarcz R. Indole-3-propionate: a potent hydroxyl radical scavenger in rat brain. Brain research. 1999;815(2):382–388.

27. Hardeland R, Zsizsik B, Poeggeler B, Fuhrberg B, Holst S, Coto-Montes A. Indole-3-pyruvic and-propionic acids, kynurenic acid, and related metabolites as luminophores and free-radical scavengers.Tryptophan, Serotonin, and Melatonin: Springer; 1999:389–395.

28. Karbownik M, Stasiak M, Zygmunt A, Zasada K, Lewiński A. Protective effects of melatonin and indole□3□propionic acid against lipid peroxidation, caused by potassium bromate in the rat kidney. Cell Biochemistry and Function: Cellular biochemistry and its modulation by active agents or disease. 2006;24(6):483–489.

29. Karbownik M, Stasiak M, Zasada K, Zygmunt A, Lewinski A. Comparison of potential protective effects of melatonin, indole□3□propionic acid, and propylthiouracil against lipid peroxidation caused by potassium bromate in the thyroid gland. Journal of cellular biochemistry. 2005;95(1):131–138.

30. Cason CA, Dolan KT, Sharma G, Tao M, Kulkarni R, Helenowski IB, Doane BM, Avram MJ, McDermott MM, Chang EB, Ozaki CK, Ho KJ. Plasma microbiome-modulated indole- and phenyl-derived metabolites associate with advanced atherosclerosis and postoperative outcomes. J Vasc Surg. 2018;68(5):1552–1562 e1557.

31. de Mello VD, Paananen J, Lindstrom J, Lankinen MA, Shi L, Kuusisto J, Pihlajamaki J, Auriola S, Lehtonen M, Rolandsson O, Bergdahl IA, Nordin E, Ilanne-Parikka P, Keinanen-Kiukaanniemi S, Landberg R, Eriksson JG, Tuomilehto J, Hanhineva K, Uusitupa M. Indolepropionic acid and novel lipid metabolites are associated with a lower risk of type 2 diabetes in the Finnish Diabetes Prevention Study. Sci Rep. 2017;7:46337.

32. Tuomainen M, Lindstrom J, Lehtonen M, Auriola S, Pihlajamaki J, Peltonen M, Tuomilehto J, Uusitupa M, de Mello VD, Hanhineva K. Associations of serum indolepropionic acid, a gut microbiota metabolite, with type 2 diabetes and low-grade inflammation in high-risk individuals. Nutr Diabetes. 2018;8(1):35.

33. Dodd D, Spitzer MH, Van Treuren W, Merrill BD, Hryckowian AJ, Higginbottom SK, L. A, Cowan TM, Nolan GP, Fischbach MA, Sonnenburg JL. A gut bacterial pathway metabolizes aromatic amino acids into nine circulating metabolites. Nature. 2017;551(7682):648–652.

34. Menni C, Hernandez MM, Vital M, Mohney RP, Spector TD, Valdes AM. Circulating levels of the anti-oxidant indoleproprionic acid are associated with higher gut microbiome diversity. Gut Microbes. 2019;10(6):688–695.

35. Koh A, De Vadder F, Kovatcheva-Datchary P, Backhed F. From Dietary Fiber to Host Physiology: Short-Chain Fatty Acids as Key Bacterial Metabolites. Cell. 2016;165(6):1332–1345.

36. De Souza CB, Jonathan M, Saad SMI, Schols HA, Venema K. Degradation of fibres from fruit by-products allows selective modulation of the gut bacteria in an in vitro model of the proximal colon. Journal of Functional Foods. 2019;57:275–285.

37. Nakata T, Kyoui D, Takahashi H, Kimura B, Kuda T. Inhibitory effects of laminaran and alginate on production of putrefactive compounds from soy protein by intestinal microbiota in vitro and in rats. Carbohydr Polym. 2016;143:61–69.

38. Ferrell M, Bazeley P, Wang Z, Levison BS, Li XS, Jia X, Krauss RM, Knight R, Lusis AJ, Garcia-Garcia JC, Hazen SL, Tang WHW. Fecal Microbiome Composition Does Not Predict Diet-Induced TMAO Production in Healthy Adults. J Am Heart Assoc. 2021;10(21):e021934.

39. Markowiak-Kopec P, Slizewska K. The Effect of Probiotics on the Production of Short-Chain Fatty Acids by Human Intestinal Microbiome. Nutrients. 2020;12(4).

40. Durazzi F, Sala C, Castellani G, Manfreda G, Remondini D, De Cesare A. Comparison between 16S rRNA and shotgun sequencing data for the taxonomic characterization of the gut microbiota. Sci Rep. 2021;11(1):3030.

