## Supplemental figures and description for "Tryptophan metabolism, gut microbiota, and carotid artery plaque in women with and without HIV infection"

**Supplementary Figures**

**Figure legends**

**Supplementary Fig1**. Sample sizes by association analysis in the present study.

**Supplementary Fig2**. Partial Spearman correlation matrix among the 10 tryptophan metabolites.

**Supplementary Fig3**. Distribution of tryptophan metabolites by HIV serostatus.

**Supplementary Fig4**. Associations of IPA levels with bacteria in the families and genera that were not captured in the 16S rRNA sequencing data but were in the same *Clostridiales* order in both 16S rRNA and shotgun sequencing data.

**Supplementary Fig5**. The associations of KEGG Ortholog groups and plasma indole-3-propionate levels.

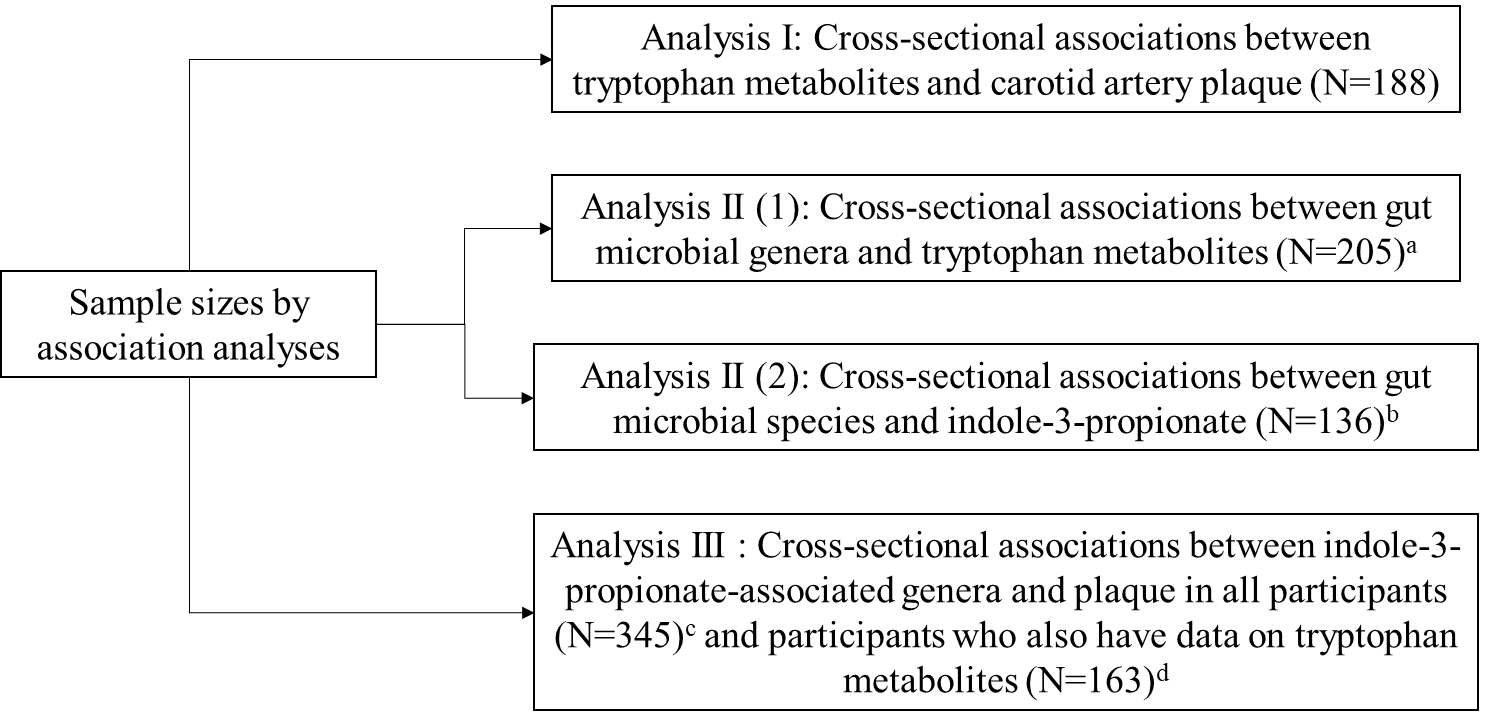

**Supplementary Fig1**. Sample sizes by association analysis in the present study. There were 361 participants with available data on both 16S rRNA sequencing and carotid artery plaque, and 251 participants had plasma tryptophan metabolites measured. Participants who took antibiotics within the four weeks of stool sample collection were removed away from the microbiome analyses. ^a^ the original number was 212, including 7 participants who took antibiotics; ^b^ Metagenomic data were available among 141 women who had tryptophan metabolites data, including 5 participants who took antibiotics; ^c^ the original number was 361, including 16 participants who took antibiotics; ^d^ the original number was 169, with 6 participants who took antibiotics.

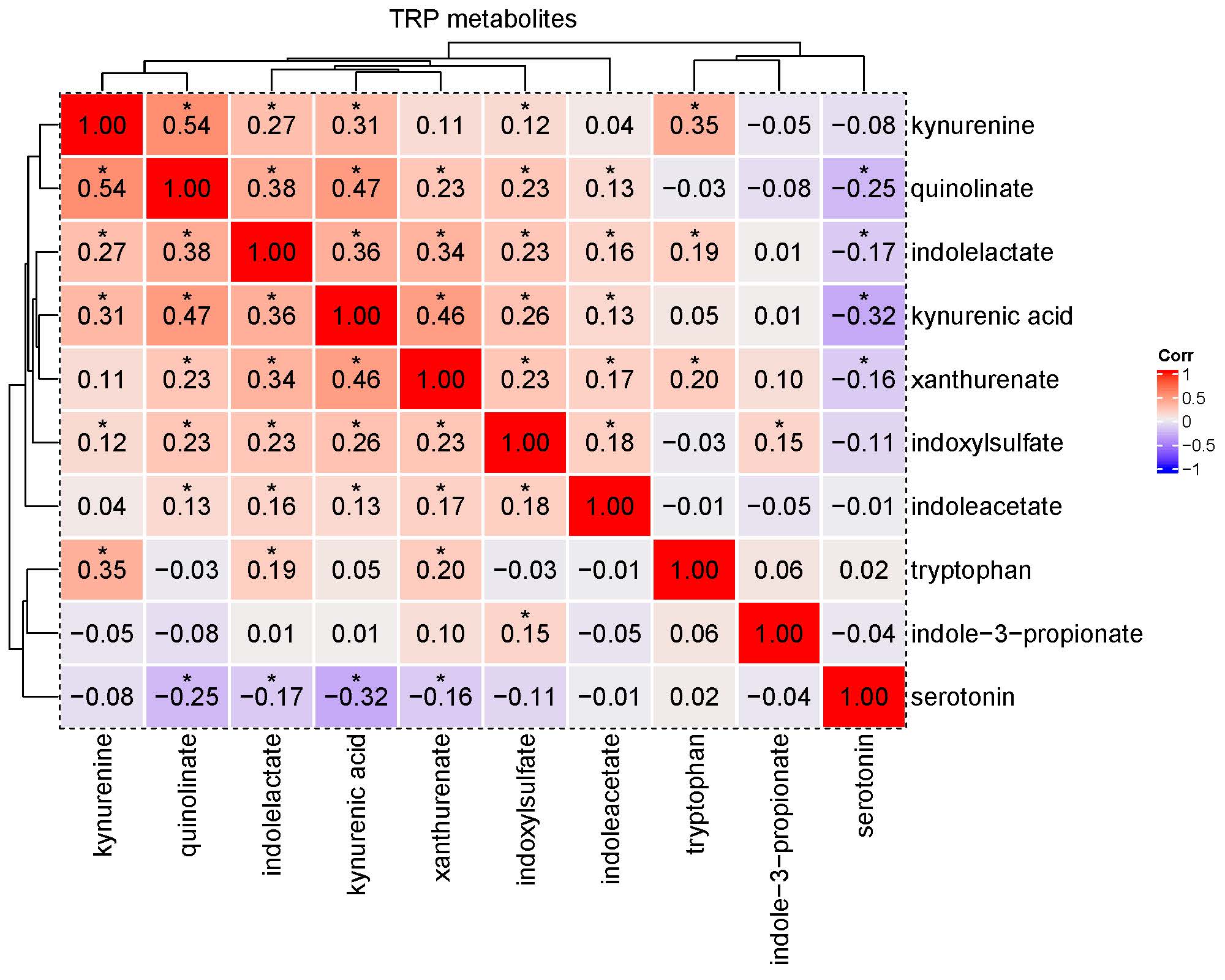

**Supplementary Fig2**. Partial Spearman correlation matrix among the 10 tryptophan metabolites. Correlation coefficients were generated after adjusting for age at visit, race/ethnicity, study sites and HIV serostatus.

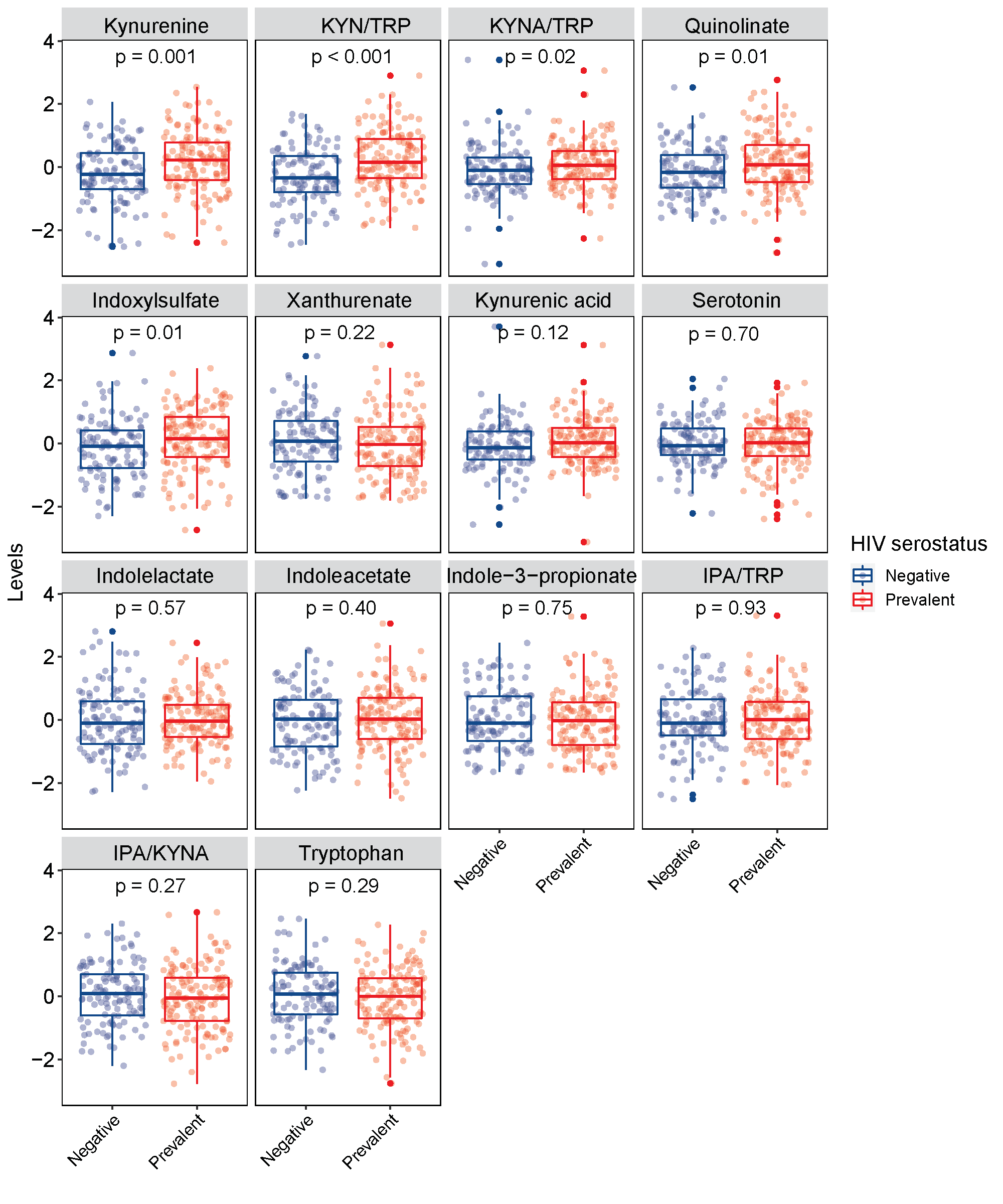

**Supplementary Fig3**. Distribution of tryptophan metabolites and constructed ratios by HIV serostatus (n=251). *P* values were derived from Wilcoxon tests based on residuals of tryptophan metabolites after adjusting for age, race/ethnicity, study sites. TRP: tryptophan; KYN: kynurenine; KYNA: kynurenic acid; IPA: indole-3-propionate.

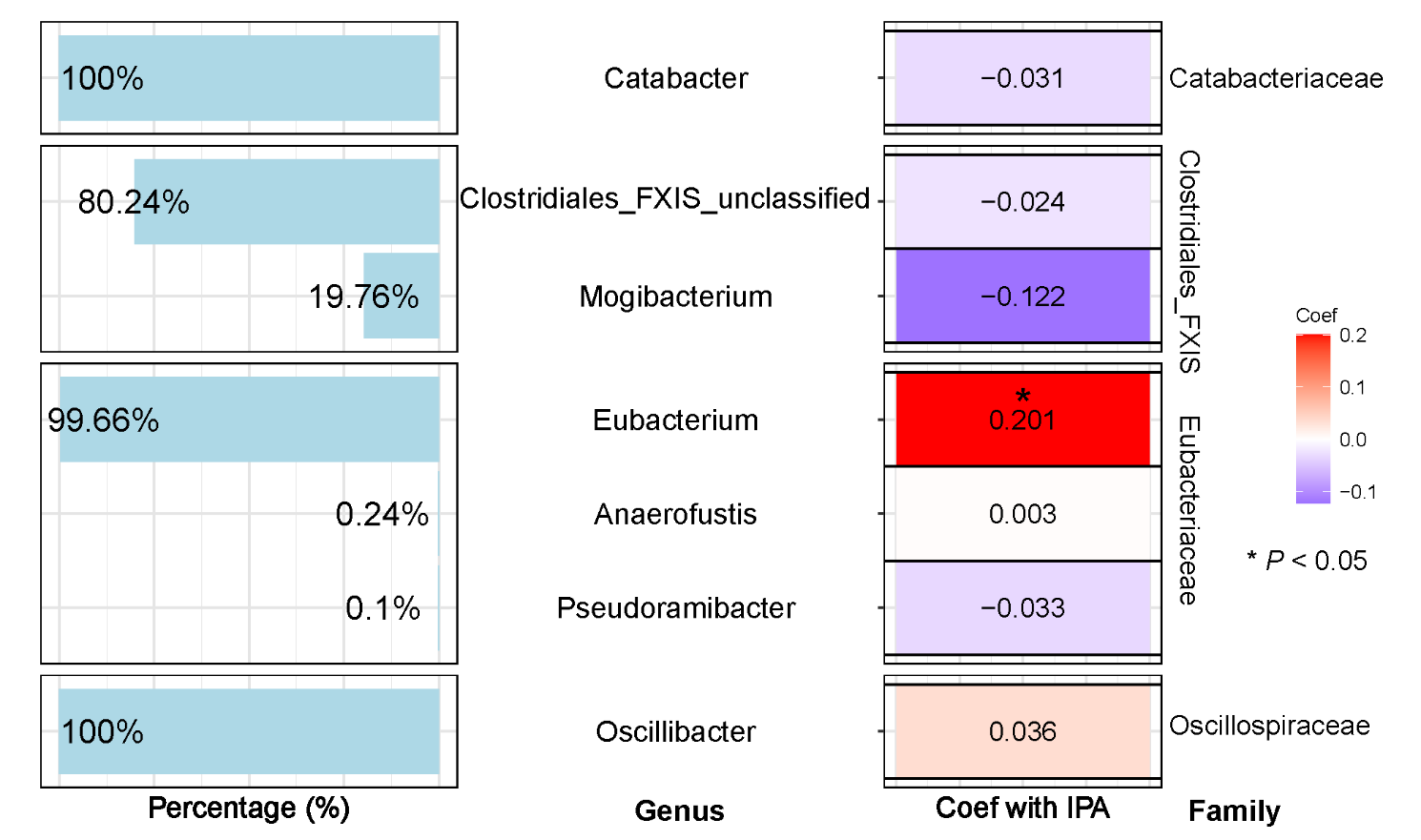

**Supplementary Fig4**. Associations of IPA levels with bacteria in the families and genera that were not captured in the 16S rRNA sequencing data but were in the same Clostridiales order in both 16S rRNA and shotgun sequencing data (n=136). Abundances of identified genera were central-log ratio (CLR) transformed. Coefficients (Coef) with IPA were adjusted for age, race/ethnicity, study sites, education, smoking, and HIV serostatus. Only taxa that were presented in more than 10% of samples with a 0.01% or higher relative abundance were included in the analysis.

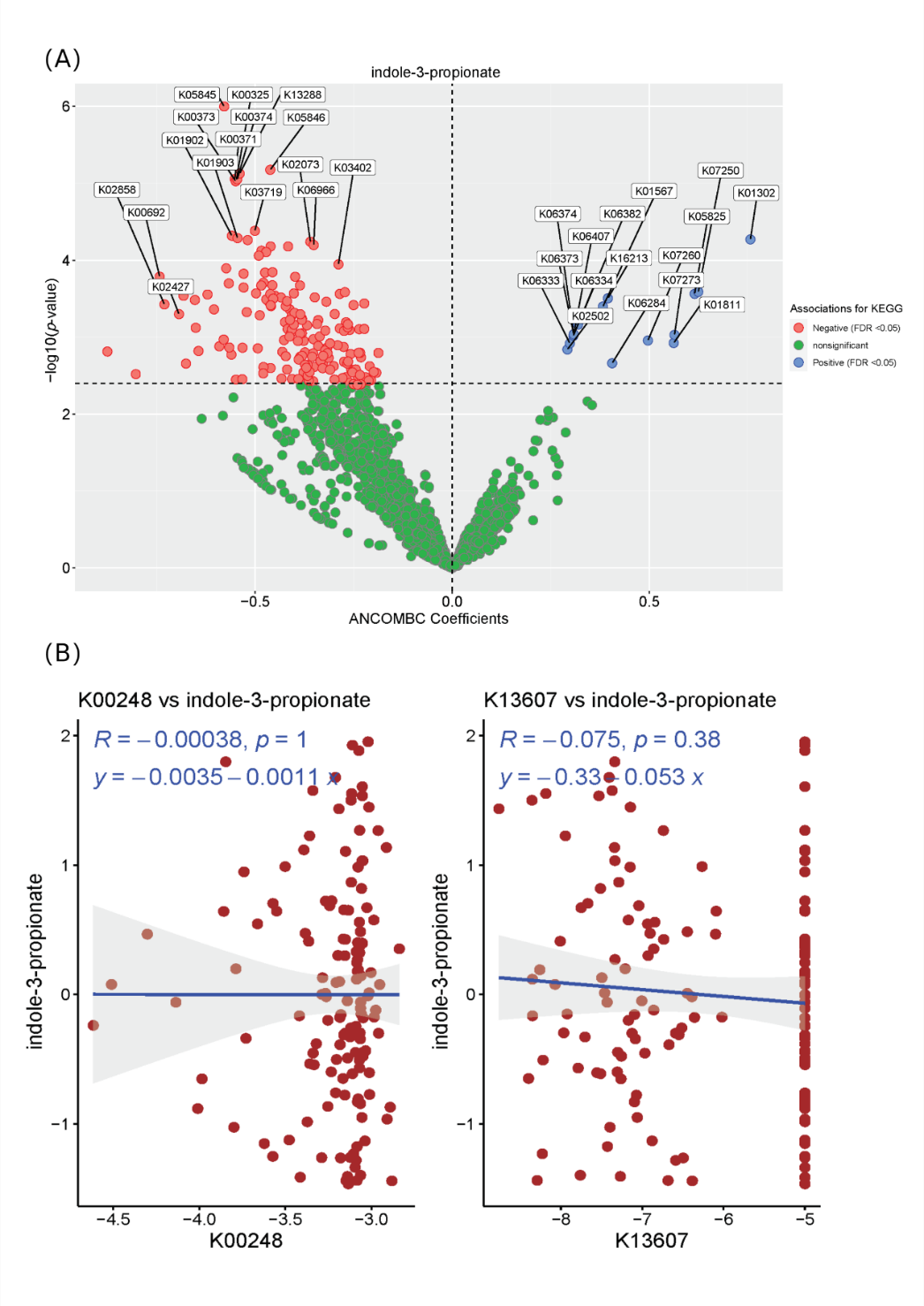

**Supplementary Fig5**. The associations of KEGG Ortholog groups (KOs) and plasma indole-3-propionate (IPA) levels (N=136). (A) volcano plot for the relationship between KOs (n=2261) and plasma IPA levels in the Analysis of Compositions of Microbiomes with Bias Correction (ANCOMBC) while adjusting for age, race/ethnicity, study sites, education level, smoking status, and HIV serostatus at visit. Red and blue dots refer to the 189 KOs having significant negative (n=173) and positive (n=16) associations with IPA, respectively. (B) the associations of two enzymes (KOs: K13607 and K00248) that were biologically involved in the tryptophan🡪 IPA pathway with plasma IPA levels among participants with shotgun sequencing data. K13607: Cs-FLDA(3-(aryl)acryloyl-CoA:(R)-3-(aryl)lactate CoA-transferase); K00248: ACDS(butyryl-CoA dehydrogenase). The enzymes abundances were central log ratio transformed, while IPA levels were inverse normal transformed. Estimates were derived through multiple linear regression with the same set of covariates in (A) being adjusted.
